# Excess COVID-19-associated deaths among the unvaccinated population ≥18 years old in the United States, May 30 – December 4, 2021

**DOI:** 10.1101/2022.02.10.22270823

**Authors:** Katherine M. Jia, William P. Hanage, Marc Lipsitch, David L. Swerdlow

## Abstract

Vaccines against SARS-CoV-2 were authorized at the end of 2020 and are effective in preventing deaths; however, many persons remain unvaccinated. Using weekly publicly available CDC data of COVID-19-associated death rates by age and vaccination status from 26 US jurisdictions, we estimated the number of excess deaths that might have been averted by vaccination among unvaccinated persons ≥ 18 years old from May 30 to December 4, 2021. We subtracted the death rate in the vaccinated from rates in the unvaccinated to estimate the death rate each week that could be attributable to non-vaccination and multiplied this rate difference by the number of people in the unvaccinated group for each age group and each week, to estimate the excess mortality among the unvaccinated. Then, we extrapolated the number of deaths due to non-vaccination in the 26 jurisdictions to the whole US population using 2020 census estimates. In the 26 participating jurisdictions there were an estimated 83,400 excess deaths among the unvaccinated from May 30 to December 4, 2021. The largest number of excess deaths occurred in those 65–79 years old (n=28,900; 34.7% of total), followed by those 50-64 years old (n=25,900; 31.1%). Extrapolated to the US population we estimated approximately 135,000 excess deaths during the study period in persons ≥18 years old. Our estimates are an underestimate of all excess deaths that have occurred since vaccine became available because our analysis period was limited to May 30 to December 4, 2021, and many excess deaths occurred before and after this period. In summary, we used retrospective data to estimate the substantial number of COVID-19-associated deaths among the unvaccinated illustrating the importance of vaccination to prevent further unnecessary mortality during this pandemic.

## TEXT

Vaccines against SARS-CoV-2 were authorized at the end of 2020 and are effective in preventing deaths; however, many persons remain unvaccinated. Using weekly publicly available US Centers for Disease Control and Prevention (CDC) data of COVID-19-associated death rates by age and vaccination status from 26 US jurisdictions (**Appendix I**), we estimated the number of excess deaths that might have been averted by vaccination among unvaccinated persons ≥ 18 years old from May 30 to December 4, 2021. May 30 was chosen because vaccine was readily available to the public by then. We subtracted the death rate in the vaccinated from rates in the unvaccinated to estimate the death rate each week that could be attributable to non-vaccination, obtained from [1] (**Appendix II**). We multiplied this rate difference by the number of people in the unvaccinated group for each age group and each week, to estimate the excess mortality among the unvaccinated. This accounted for the changing attack rate, and changing proportion vaccinated, during this time period. Then, we extrapolated the number of deaths due to non-vaccination in the 26 jurisdictions to the whole US population using 2020 census estimates, after excluding those partially vaccinated [2] (**Appendix III**).

The 26 US jurisdictions had similar age structure to the overall US population and a similar cumulative incidence of COVID-19 per 100 persons. However, the 26 jurisdictions had a higher proportion unvaccinated compared to that in the US overall, when considering only the fully vaccinated and unvaccinated (**Table 1**).

**Table 1.** Comparing the 26 participating jurisdictions with the overall US population

|  | <u>26 jurisdictions</u><br>(from ref [1]) | <u>US</u> | <u>Source(s) of data for US</u> |
| --- | --- | --- | --- |
| **Population** |  |  |  |
| Number of individuals aged 18 years old and above (mean) | 146,611,787<sup>a</sup> | 256,662,010<sup>b</sup> | [2] |
| **Age distribution (%)** |  |  |  |
| 18-29 | 21.2 | 20.8 |  |
| 30-49 | 33.2 | 33.1 |  |
| 50-64 | 24.4 | 24.5 |  |
| 65-79 | 16.1 | 16.6 |  |
| 80+ | 5.1 | 5.1 |  |
| **Vaccination** |  |  |  |
| % unvaccinated / (fully vaccinated + unvaccinated)<sup>c</sup> | 33.9% | 28.1% | [3] |
| **COVID-19 impacts, May 30 – December 4, 2021** |  |  |  |
| Cumulative COVID-19 incidence | 4.6 per 100 persons<sup>d</sup> | 4.7 per 100 persons<sup>e</sup> | [4, 5] |
| Total number of COVID-19 associated deaths | 111,629<sup>f</sup> | 196,447<sup>g</sup> | [4] |
| **Estimated excess number of COVID-19 associated deaths among unvaccinated individuals (18 years old and above), May 30 – December 4, 2021** | 83,400 | 135,000<sup>h</sup> | [2] |
| **Age distribution of excess deaths (%)** |  |  |  |
| 18-29 | 2.6 | 2.4 |  |
| 30-49 | 14.1 | 13.9 |  |
| 50-64 | 31.1 | 30.8 |  |
| 65-79 | 34.7 | 35.4 |  |
| 80+ | 17.6 | 17.5 |  |

<sup>a</sup> Partially vaccinated individuals were excluded from the data set for the 26 jurisdictions, so only those fully vaccinated and unvaccinated individuals counted towards the total.
<sup>b</sup> From the US 2020 population estimates by year-age.
<sup>c</sup> Total number of unvaccinated divided by total number of fully vaccinated and unvaccinated, after excluding those partially vaccinated among those 18 years old and above, averaged across the study period.
<sup>d</sup> Cumulative incidence was calculated as the total number of new reported cases with a positive specimen collection date from May 30 to December 4 divided by the mean population during the period (12 years old or above) in 28 jurisdictions (the 26 jurisdictions plus North Carolina and New York state).
<sup>e</sup> This estimate was obtained by dividing the total number of new reported cases during the study period by the US population estimates in August 2021 (including those <12 years old).
<sup>f</sup> Included deaths occurring among those 18 years and above with a positive specimen collection date from May 30 to December 4, 2021.
<sup>g</sup> Included all-age COVID-19-associated deaths in the US with a date of reporting from May 30 to December 4, 2021. Note that this is not directly comparable to the figure from 26 jurisdictions (due to different age ranges and use of report date rather than test date).
<sup>h</sup> Extrapolated based on the US 2020 population estimates for 18 years old or above, under the assumption that the rest of the country had the same proportion unvaccinated as the 26 participating jurisdictions after excluding those partially vaccinated (Appendix II).

In the 26 participating jurisdictions there were an estimated 83,400 excess deaths among the unvaccinated from May 30 to December 4, 2021. The largest number of excess deaths occurred in those 65–79 years old (n=28,900; 34.7% of total), followed by those 50-64 years old (n=25,900; 31.1%) (**Appendix IV**). We extrapolated the estimate to the US population after excluding those partially vaccinated based on national data on vaccination rates [3], and estimated approximately 135,000 excess deaths during the study period in persons ≥18 years old. In our sensitivity analyses, the excess deaths varied by the assumed size of the unvaccinated population in the non-26 jurisdictions, which dropped to 119,000 were the population that was unvaccinated 30% lower, and increased to 150,000 were it 30% higher, in the rest of the country (**Appendix V**), and also varied by proportion partially vaccinated (**Appendix VI**).

We used data that represented over half of the total US population and the 26 US jurisdictions have age structure and incidence similar to the nation overall. In addition, rates of underlying conditions and prevention behaviors may vary between vaccinated and unvaccinated individuals and confer differing risks for acquisition of SARS-CoV-2 infection and mortality. Although we cannot be certain that all the excess deaths in unvaccinated could have been prevented with vaccination, subtracting rates of death among vaccinated from deaths in unvaccinated should partially account for deaths that would have occurred even if persons were vaccinated. Handling of partial vaccination is a challenge because the proportion partially vaccinated is not reported for the 26 jurisdictions and because of possible reporting lags and misclassification [3]. This was further evaluated by conducting a sensitivity analysis in **Appendix VI**. Our estimates are an underestimate of all excess deaths that have occurred since vaccine became available because our analysis period was limited to May 30 – December 4, 2021 and many excess deaths occurred before and after this period. In addition, we only estimated direct effects of vaccination, however the number of excess deaths could have been higher if indirect effects were included since vaccines likely reduce transmission to others. In summary, we used retrospective data to provide an estimate of the substantial number of COVID-19-associated deaths among the unvaccinated illustrating the importance of vaccination to prevent further unnecessary mortality during this pandemic.

## Data Availability

All data used in this publication is publicly available from the US CDC. https://covid.cdc.gov/covid-data-tracker/#rates-by-vaccine-status

## Acknowledgement

We acknowledge the CDC for curating and sharing the data for this analysis, with special thanks to Akilah Ali, Avnika Amin, Amelia Johnson, and Heather Scobie, for coordinating with individual states, establishing standardized methods, and collating the data.

## Supplementary Materials

## Appendix I

**Table S1.**
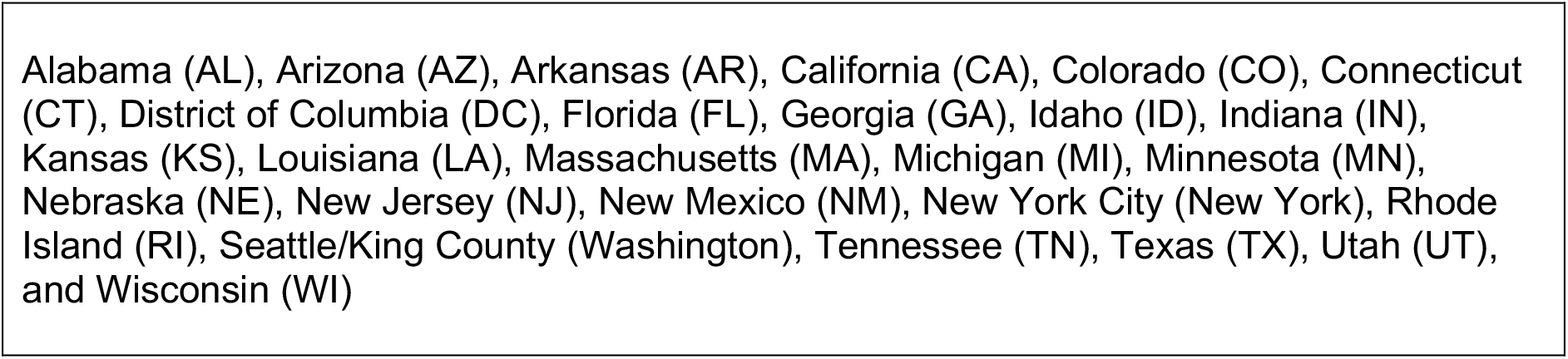
List of the 26 participating jurisdictions (version date: January 21, 2022)

## Appendix II

**Figure S1.**
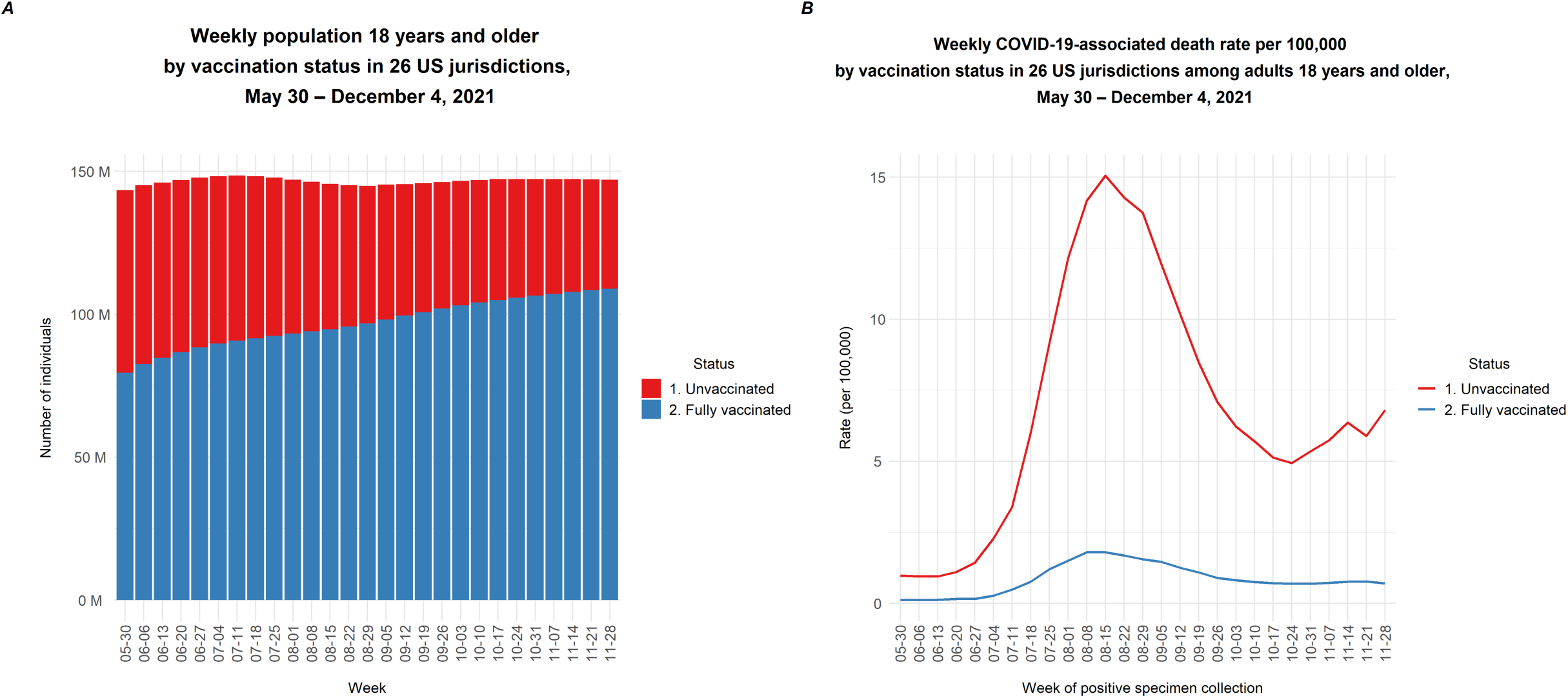
Population size and COVID-19-associated death rates among adults 18 years and older by vaccination status in 26 US jurisdictions, May 30 – December 4, 2021. The bar plot shows the total number of individuals 18 and older in the 26 jurisdictions by vaccination status (partially vaccinated individuals were excluded) (A). The line graph shows the mortality rate (per 100,000) for COVID-19-associated deaths in the two groups. The number of excess deaths was estimated by multiplying the red bars in (A) by the difference between the red and the blue lines in (B) (i.e., excess mortality rate among the unvaccinated population), by age and week.

## Appendix III Data sources and analysis

### Data on COVID-19-associated deaths by vaccination status

Data on vaccination status and COVID-19-associated deaths were from the US CDC, aggregated across participating US jurisdictions (CDC, 2022a). Included jurisdictions had to regularly link their case surveillance and immunization data with CDC. Twenty-six of the jurisdictions reported deaths among vaccinated and unvaccinated persons and were included in the analysis.

### Vaccination status

Individuals were considered as fully vaccinated if they completed a primary series of a COVID-19 vaccine authorized by the Food and Drug Administration (FDA) at least 14 days before the date of record (Scobie et al 2021). The number of unvaccinated was estimated by subtracting the numbers of fully and partially vaccinated (which were determined by vaccine administration data) from the US 2019 population estimate (Johnson et al 2021). However, the partially vaccinated individuals, who had at least one dose of the approved vaccine but did not complete the series at least 14 days before the time of record, were excluded from the data set (CDC, 2022a). Correspondingly, we excluded the partially vaccinated when extrapolating the number of excess deaths from the 26 jurisdictions to the US total.

### Ascertainment of COVID-19-associated deaths

A COVID-19-associated death was defined as a person who had a positive documented SARS-CoV-2 diagnosis and died. The data set recorded deaths among vaccinated and unvaccinated individuals based on the week of a positive specimen collection, which was usually up to 30 days ahead of the date of death (CDC, 2022a). Methods of outcome ascertainment varied across jurisdictions: it was common to use vital records, while some jurisdictions used a combination of vital records and provider reporting and/or case investigations (Scobie et al, 2021).

### Mortality rate difference

For each week in the study period, we estimated the mortality rate difference, by subtracting the rate of deaths among unvaccinated individuals with that among vaccinated individuals, for each age group in the 26 jurisdictions. This measure will account for both the changing attack rate over the course and the proportion vaccinated that increased over time.

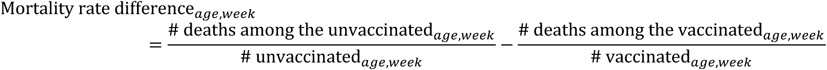

We then multiplied the mortality rate difference to the total number of unvaccinated individuals in the 26 jurisdictions by age group and week, to estimate the number of excess deaths.

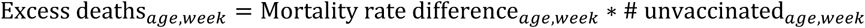

Thirdly, we extrapolated the excess deaths to the whole US population, with the partially vaccinated excluded from the numerator (see formula below). The percentage partially vaccinated was obtained from another CDC data set on vaccination demographics in the US. We subtracted the percentage fully vaccinated from the percentage vaccinated with at least 1 dose(s) by week, for each age group: the 18-24, 25-39, 40-49, 50-64, 65-74, and the 75+ years old (CDC, 2022b), to obtain the percentage partially vaccinated.

We multiplied the total US resident population of each year-age with the factor (1 - %partially vaccinated), for each age group (US Census Bureau, 2022), and then extrapolated the excess mortality as follows:

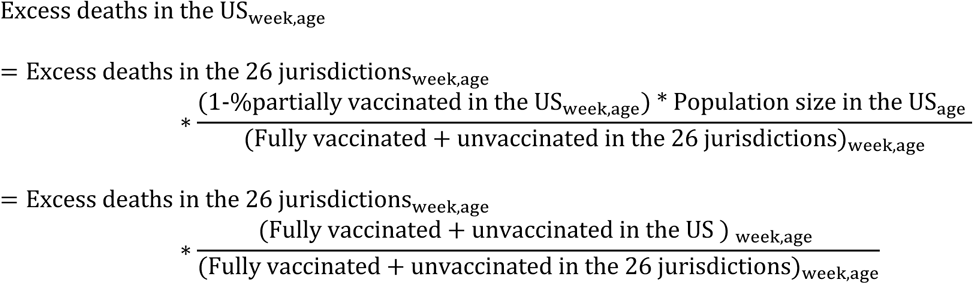

Lastly, we summed the weekly number of excess deaths by age and by week to obtain the total number of excess deaths among the unvaccinated population from May 30 to December 4, 2021.

## Appendix IV Estimated weekly excess COVID-19-associated deaths among unvaccinated populations 18 years old and above, May 30 – December 4, 2021

**Figure S2.**
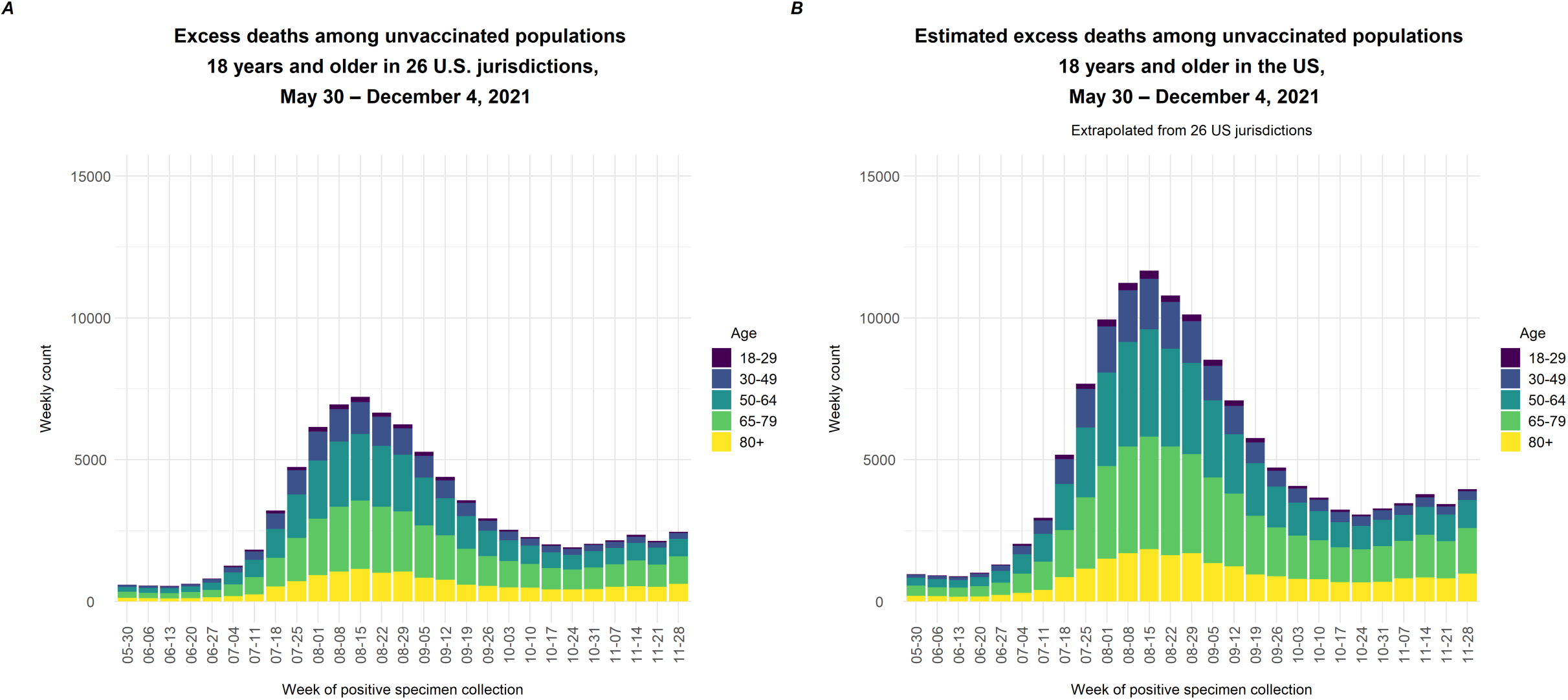
Estimated weekly excess COVID-19-associated deaths among unvaccinated populations 18 years and older in 26 US jurisdictions (A) and in the US (B), May 30 – December 4, 2021

**Table S2.**
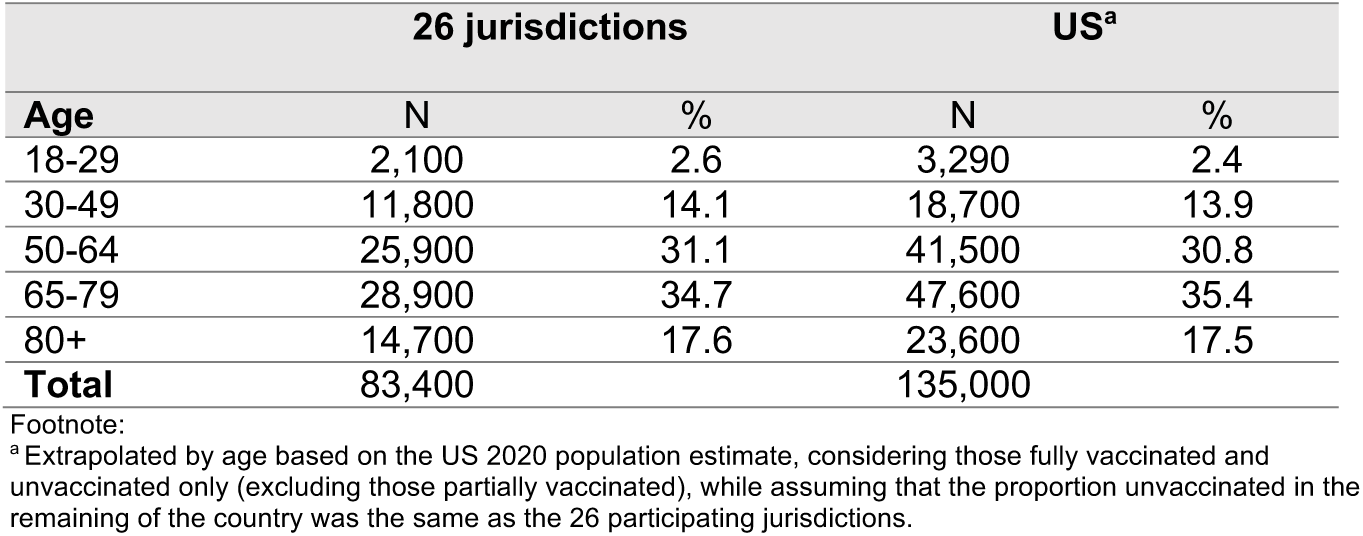
Estimated excess COVID-19-associated deaths by age group among unvaccinated populations 18 years old and above, May 30 – December 4, 2021.

## Appendix V Estimated total number of excess deaths under varying assumptions about the proportion of the population unvaccinated in the non-26 US jurisdictions, May 30 – December 4, 2021

The excess mortality for the whole US population is given by

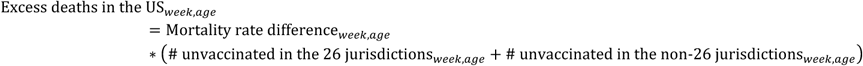

By allowing the number of unvaccinated in the non-26 jurisdictions to change by a percentage *m%*, we have

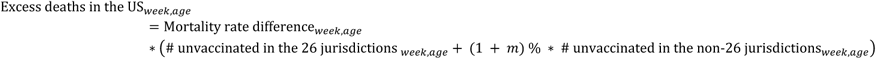

The resulting estimates are:

**Figure S3.**
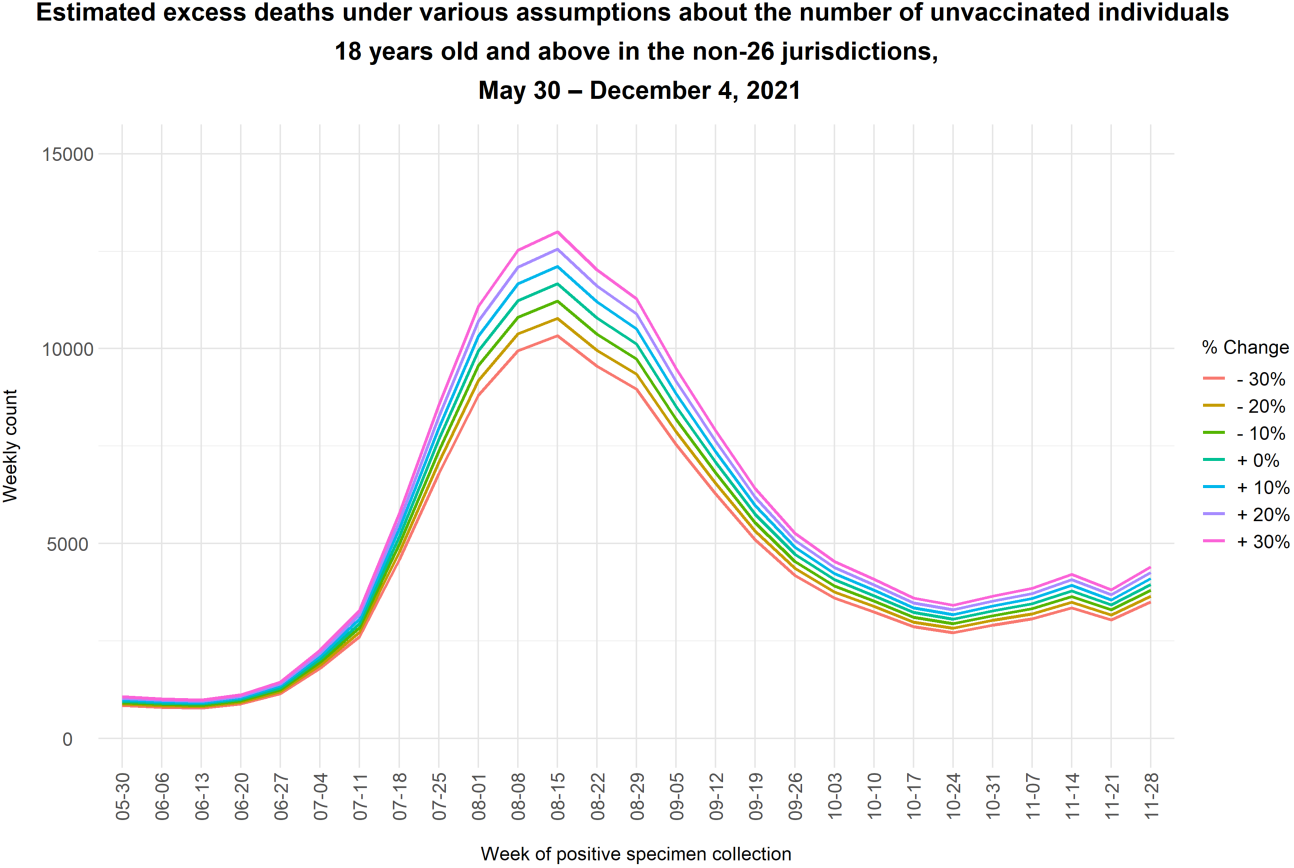
Estimated weekly number of excess deaths among the unvaccinated population in the US using different assumptions about the proportion unvaccinated in the non-26 US jurisdictions, May 30 – December 4, 2021. The numbers of fully vaccinated and unvaccinated individuals and their mortality rates were derived from the data set for 26 participating US jurisdictions, and therefore the total number of excess deaths in those 26 jurisdictions was unvaried in this sensitivity analysis.

**Table S3.**
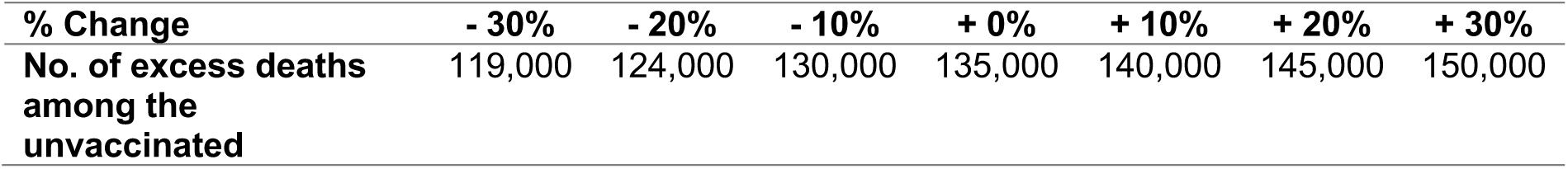
Estimated total number of excess deaths among the population 18 and above in the US, May 30 – December 4, 2021, under different assumptions about the proportion unvaccinated in the non-26 US jurisdictions, after excluding those partially vaccinated.

## Appendix VI Estimated total number of excess deaths under varying assumptions about the proportion partially vaccinated in the United States, May 30 – December 4, 2021

In this sensitivity analysis, we extrapolated the excess deaths among the unvaccinated by considering different percentage changes in the proportion partially vaccinated in the whole US population. We allow it to change by *p%* for each scenario, as in:

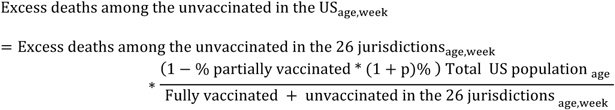

As seen from the plot below, the estimated excess deaths decreased if there were a higher percentage of partially vaccinated in the population, and vice versa.

**Figure S4.**
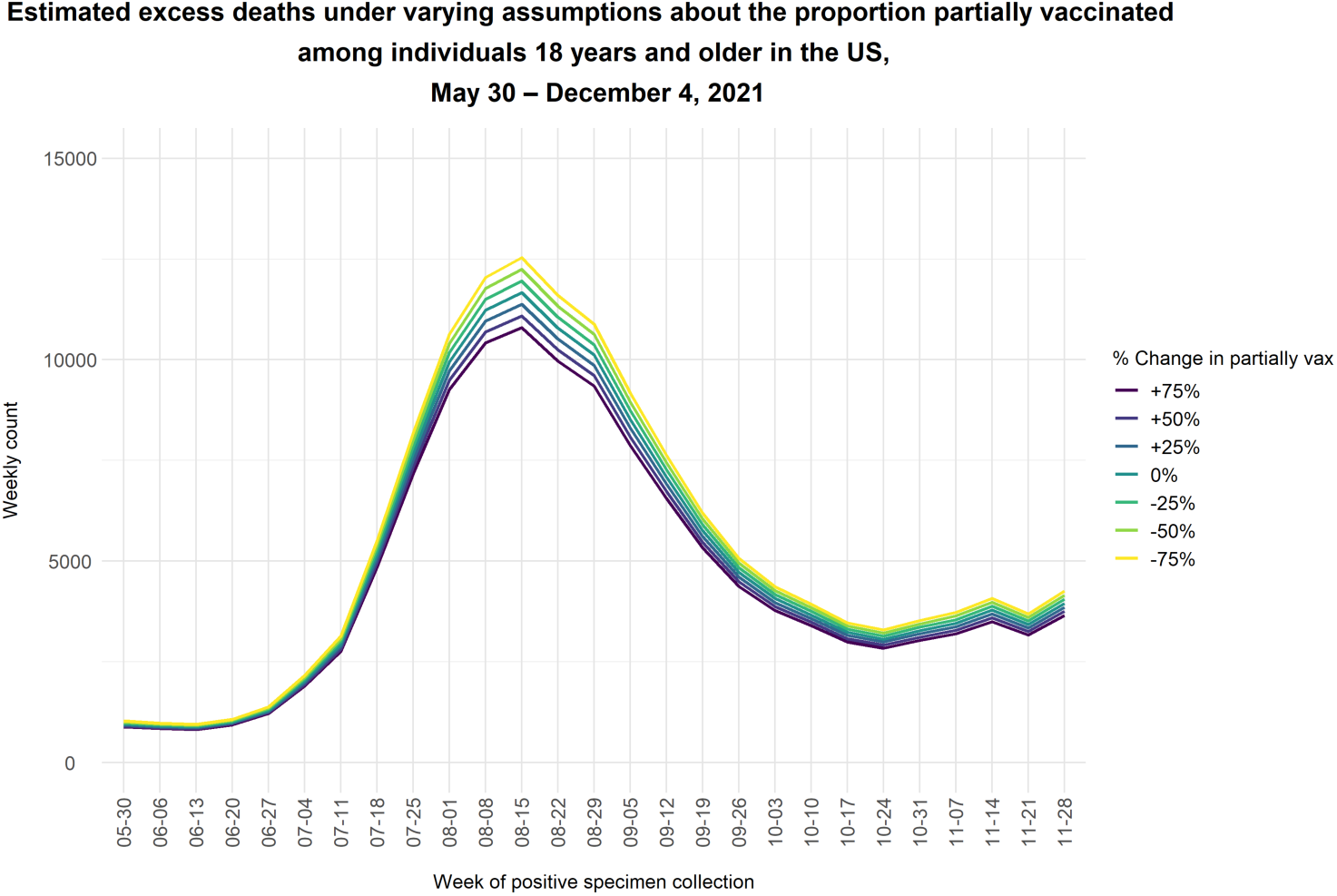
Estimated excess deaths under varying assumptions about the proportion partially vaccinated among individuals 18 years and older in the US, May 30 – December 4, 2021

**Table S4.** Estimated total number of excess deaths among those 18 years old and above in the US, May 30 - December 4, 2021, by varying the proportion partially vaccinated in the population

| Change in the % partially vaccinated in the population | -75% | -50% | -25% | 0% | +25% | +50% | +75% |
| --- | --- | --- | --- | --- | --- | --- | --- |
| No. of excess deaths among the unvaccinated | 144,000 | 141,000 | 138,000 | 135,000 | 131,000 | 128,000 | 125,000 |

